# Towards Non-Invasive White Blood Cell Count in Humans

**DOI:** 10.1101/2024.11.26.24314245

**Authors:** Arutyun Bagramyan, Juwell W. Wu, Kamdin Mirsanaye, Clemens Alt, Charles P. Lin

**Affiliations:** Center for Systems Biology and Wellman Center for Photomedicine, Massachusetts General Hospital and Harvard Medical School, Boston, MA

**Author notes:** **Materials & Correspondence** Correspondence and requests for materials should be addressed to A.B.

## Abstract

Despite rapid advances in diagnostic and imaging technologies, a method for noninvasive monitoring of the immune system does not exist. The standard white blood cell count (WBCC), a key clinical measure for assessing patients’ health, requires drawing blood, which poses inherent risks for secondary infection and anemia in vulnerable patient populations. In addition, the specialized equipment, expertise, and infrastructure are not always available in resource-poor settings. Here we present a method for noninvasive and label-free WBCC by imaging human oral mucosa with a miniaturized oblique back-illumination microscope (mOBM). In a pilot study involving 34 healthy subjects, we validated our system’s ability to detect and quantify circulating leukocytes and compared our image-based WBCC to standard laboratory measurements. The ability to perform noninvasive WBCC will enable real-time assessment of the immune status during infection and inflammation or in response to therapeutic intervention without repetitive blood sampling.

## Introduction

The white blood cell count (WBCC) is a cornerstone in clinical practice, widely employed in preventive and emergency medicine to diagnose infection, inflammation, and monitor response to treatment^1^. Standard blood test requires phlebotomy, which can be challenging in vulnerable patient populations, as it poses a risk of infection in immune-compromised patients or anemia in preterm infants^2^. A method for noninvasive measurement of WBCC could alleviate phlebotomy-related complications and enable safe immune status assessments in vulnerable patient populations. Another key application lies in the preventive monitoring of healthy populations, where the normal but elevated leukocyte counts have been linked to disguised systemic inflammation associated with disease development, particularly in elderly and high-risk populations prone to developing cancer, diabetes (type 2), and cardiovascular diseases, among others^3^. The frequent monitoring of immune status with mOBM could enable early detection and treatment of such subclinical conditions.

The development of noninvasive methods for obtaining WBCC has been hindered by the absence of suitable instrumentation that can access patient vasculature, accurately resolve circulating white blood cells (WBCs) in rapidly flowing blood, and record data for a sufficient duration to ensure reliable measurements, as well as the lack of analytic tools for objective cell detection and quantification. Commercially available hand-held vital microscopes can monitor microvascular perfusion but do not provide a contrast mechanism for delineating individual white blood cells in the circulation^4,5^. Reflectance confocal microscopy^6–8^ and spectrally encoded confocal microscopy/flow cytometry^9^ are capable of high-resolution cellular imaging *in vivo*, but the presence of speckles compromises the image quality. Moreover, the strong backscattering from both red and white blood cells results in poor contrast between the two cell types when intermixed in the flowing bloodstream^8,9^. Nonlinear optical techniques such as third-harmonic generation^10^ and two-photon-induced autofluorescence microscopy^11^ have been explored, but their complexity and high instrumentation costs pose a formidable barrier to clinical translation.

## Results

Here we have developed a miniaturized oblique back-illumination microscope (mOBM) for real-time imaging and quantification of circulating leukocytes in the microvasculature of the human oral mucosa (Fig. 1a-c, Supplementary Fig. 1). We chose the lower lip as the imaging site for its favorable characteristics^9,12^, including the large perfusion bed (Fig. 1b-c), thin stratum corneum layer, low pigmentation compatible with individuals with dark skin color, comfort for the subjects, and the ease for exposing and stabilizing tissue while minimizing motion artifacts during imaging (*Materials and Methods*). For imaging, we employ the oblique back illumination (OBM) approach that generates phase-gradient contrast in thick tissue^13–15^, producing clear delineation of cell borders, which is essential for detecting and distinguishing individual white blood cells (WBC) from plasma gaps (Fig. 1d, i-v, Supplementary Video 1 and 2). WBCs that travel together as doublets or triplets can also be visualized (Fig. 1d, vi). Additional contrast is generated by the 565nm illumination wavelength that is strongly absorbed by the RBCs^14^, rendering them darker and readily distinguishable from the much brighter WBCs (Fig. 1d, f). Other key features of the mOBM are the short exposure time (≤0.5ms) that reduces motion blur and the high acquisition frame rates (up to 300 fps) that enable frame-to-frame detection of rapidly moving cells in the bloodstream (Fig. 1f).

**Figure 1.**
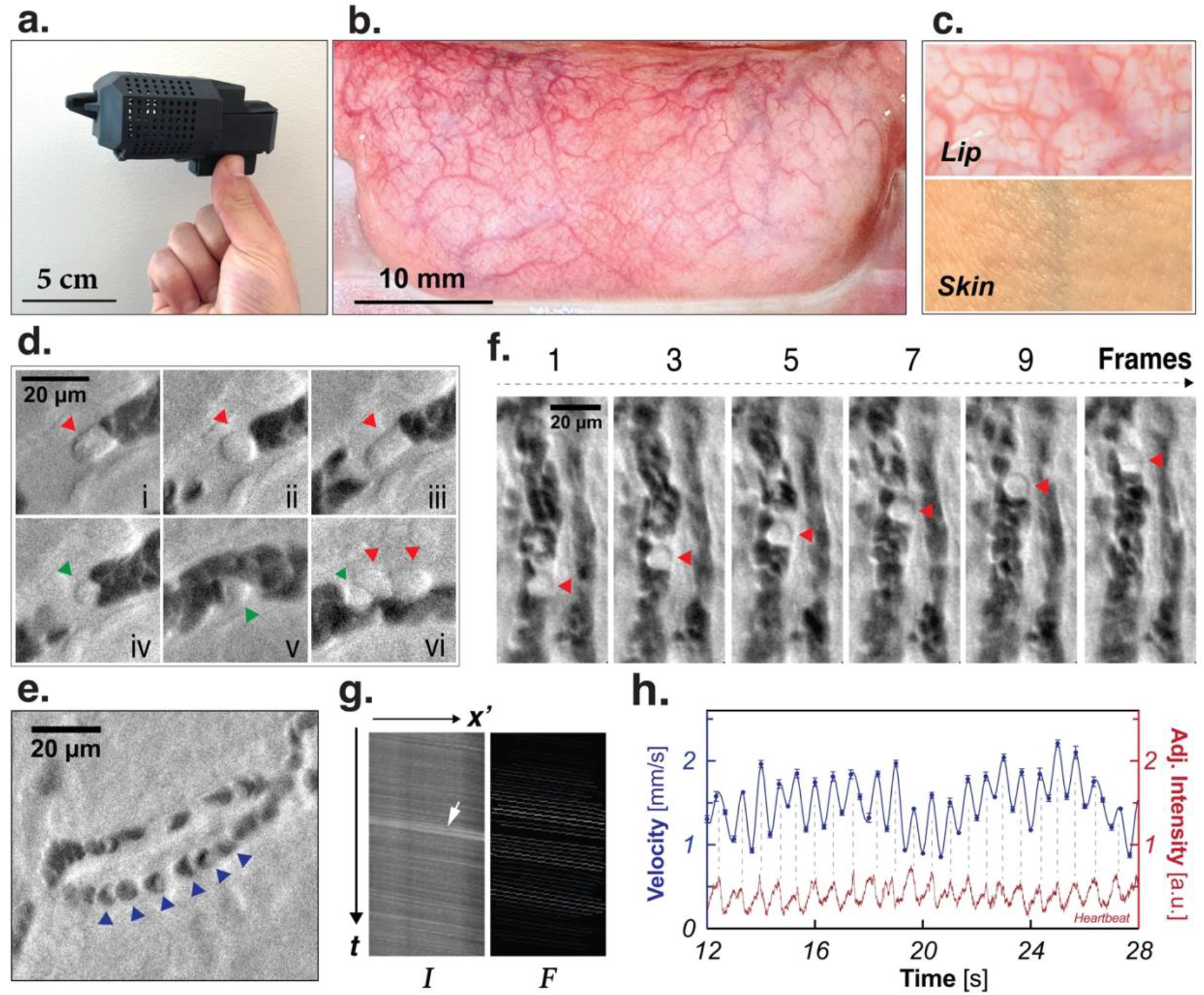
**a**. The miniaturized oblique back illumination microscope (mOBM). **b**. Exposed vasculature bed of the lower lip. **c**. Improved visualization of the vasculature in the lip compared to the skin tissue. **d**. Examples of leukocytes in the microvasculature imaged with the mOBM. Large cells (≥10 µm), most likely neutrophils, are indicated with red arrowheads (i-iii). Small leukocytes (≤10 µm), most likely lymphocytes, are indicated with green arrowheads (iv-vi). **e**. Individual RBCs (blue arrowheads) in the capillary flow. **f**. A sequence of images of a rapidly flowing leukocyte (red arrowheads), acquired at 250fps. **g**. Space-time diagram reconstructed using the pixel intensity along flow direction as a function of time across 200 consecutive images (*Materials and Methods*); I: Initial (pre-processing) F: Final (after processing). **h**. Example of a velocity and heartbeat measurement as a function of time.

To measure blood flow velocity in a user-defined region-of-interest (ROI), our image processing pipeline (*Materials and Methods*, and *Supplementary Methods*) plots the pixel intensity along the flow direction as a function of time to generate space-time diagrams, wherein the slopes are the inverse of the flow velocity values (Fig. 1g-h). In a subset of blood vessels, presumably arterioles, the velocity is found to fluctuate in sync with the heartbeat (Fig. 1h). The blood volume rate can then be obtained by multiplying the velocity by the vessel’s cross-sectional area. WBCC is defined as the number of WBCs per unit volume.

To count WBCs, our image processing pipeline automatically defines a cell count window within the ROI (Fig. 2a). Because of the differential absorption between the WBCs and RBCs, the pixels’ brightness within the window increases when a WBC passes by. Figure 2b shows an example of a temporal trace obtained by plotting the mean pixel intensity within the window as a function of time after background correction. Peak positions indicate the video frames at which leukocytes appear in the window. The pipeline then automatically counts the peaks whose intensity surpasses a moving threshold (Fig. 2b, dotted gray line). To assess the accuracy of the automated peak detection, we compared the results with cell counts from manual counting and found good agreement between the measurements (Fig. 2c, ratio=0.96±0.11). Note that manual cell counting is only possible for blood vessel diameters that are smaller than ∼22 µm. In larger vessels, some leukocytes may be out of focus in the flow stream, and leukocytes in deeper parts of the blood vessel can be obscured by RBCs at shallower depths.

**Figure 2.**
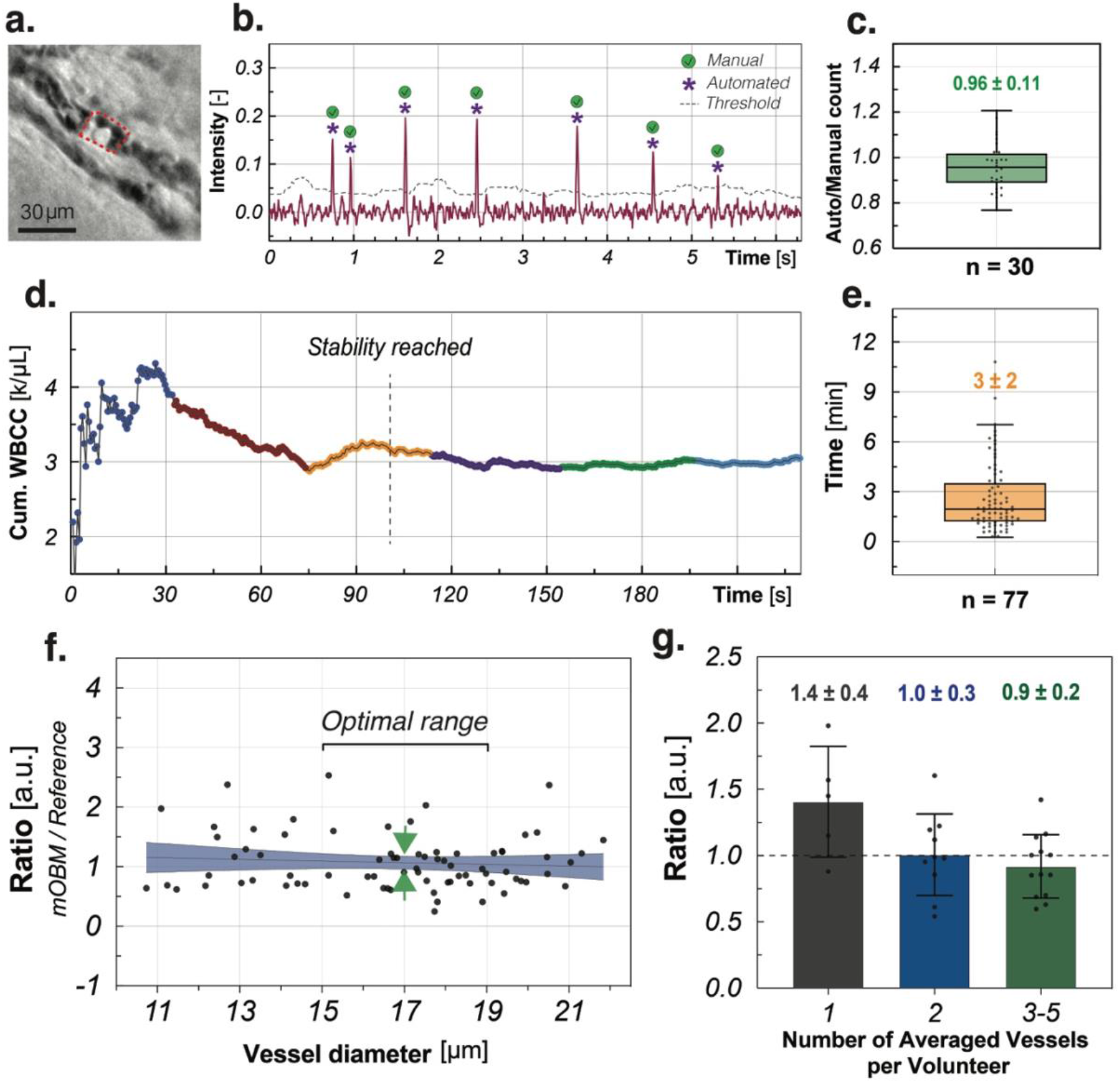
**a-c**. Method for counting circulating leukocytes. The mean of pixel values in the cell count window marked by the red dotted line in ***a***) is displayed over time as shown in ***b***). The peaks that exceed the time-dependent threshold value (dotted gray line) are counted by the automated pipeline as WBCs (*Materials and Methods*). The purple stars indicate the peaks identified by the automated pipeline, and the green checkmarks indicate the manually identified WBCs in the corresponding video. **c**. The ratios of automatic over manual cell counts of the same video (*Materials and Methods*). **d**. Example of cumulative WBCC as a function of acquisition time for a single vessel, which fulfilled our stability criterion (<5% fluctuation) in ≈101 seconds. **e**. The distribution of stabilization time from 77 vessels and 28 subjects. Each data point represents a single vessel. **f**. The ratio between the corrected WBCC obtained by our imaging method divided by the reference value obtained by drawing blood. Each point is an individual measurement from a single blood vessel. The 95% confidence interval (CI) of a linear fit is indicated with the blue color. The green arrows indicate the narrowest CI area, suggesting the optimal vessel diameter to be centered at ∼17 µm. **g**. Ratio variations when averaging measurements from multiple vessels from the same subject. Each data point is from a single subject. Subjects with stabilized counts in only a single blood vessel are represented by the gray bar, while subjects with stabilized counts in 2 or 3-5 vessels are represented by blue, and green bars, respectively. The error bars represent the standard deviation (SD). The mean and SD are indicated at the top of the box plots.

Next, we used mOBM to image the microvasculature in a cohort of 34 healthy volunteers and compared our image-based WBCC to existing clinical standards obtained by drawing blood from the same subjects after imaging. We analyzed 123 blood vessels to investigate the minimum blood volume (and hence the acquisition time, or the length of video) needed to obtain a reliable WBCC measurement. As shown in Fig. 2d, the cumulative WBCC fluctuates initially but eventually stabilizes as the imaging time increases. Of the 123 vessels analyzed, 77 fulfilled the stability criterion (see *Materials and Methods)*. The acquisition time needed for stabilized WBCC measurement is 3±2 min (Fig. 2e), corresponding to a sampling volume of 61±59 nL, depending on the vessel size and flow velocity.

We evaluate the ratio R between the WBCC obtained from mOBM and from the standard lab test after blood draw (R=1 if the two measurements are in perfect agreement). As shown in Supplementary Fig. 2, R obtained from individual vessels depends on vessel size, decreasing with increasing vessel diameter, as a greater fraction of WBCs is either out of focus or masked by more superficial RBCs. We apply a correction factor to account for the undercounting at larger vessel diameters (see *Materials and Methods*). The result after the correction is shown in Figure 2f. Interestingly, the confidence interval (CI) of the linear fit presented in Figure 2f is narrower for 15-19 µm diameter vessels, suggesting that this size range may be preferable for most consistent WBCC measurements. Coincidentally, those are also the most abundant vessel sizes in the superficial oral mucosa.

Finally, we report the WBCC obtained from mOBM as the average count from multiple blood vessels within the same subject. Averaging measurements from multiple vessels should naturally lead to the reference WBCC value since phlebotomy samples blood from large veins that drain numerous microvessels. As shown in Figure 2g, the ratio starts to converge and stays close to 1 as the number of averaged vessels increases. Importantly, the mean absolute percentage error (MAPE) on WBCC measurement decreases from 45% to 27% when imaging 3-5 vessels compared to a single vessel, leading to more precise measurements that align closer with the reference WBCC.

## Discussion

We present a miniaturized microscope system for non-invasive and label-free monitoring of the systemic white blood cell count (WBCC) - a hallmark of inflammation and a key clinical indicator of patient health.

To validate our system, we imaged 34 healthy volunteers, demonstrating the ability to acquire high-quality videos across a wide spectrum of skin colors, ages, and sexes. By acquiring temporal recordings from various vessels of the oral microvasculature and assessing the WBCC over time, we showed that after initial fluctuations, measurement stabilizes in most of the vessels (≈63%). Using data from 77 stabilized vessels, we established the first benchmarks for stabilized image-based WBCC measurement, indicating an average acquisition time of 3±2 minutes, corresponding to a sampling volume of 61±59 nL per vessel. The ability to perform extended temporal recordings is crucial for reliable image-based WBCC measurement and represents a key distinction between our mOBM system and existing commercially available handheld microscopes^4,5^, which are often limited to short video acquisitions owing to severe motion and pressure artifacts.

The analysis of stabilized WBCCs showed that the detection efficiency declined with increasing vessel diameter (Supplementary Figure 2). We empirically derived a vessel size-dependent correction factor and compared the corrected WBCC measurements with the clinical reference values (Fig. 2f). The results indicate that WBCC measurements from vessels with ≈15-19 µm diameters align closest with the clinical reference value (Fig. 2f, green arrowheads). Therefore, our instrument targets vessels in this size range for optimal image-based WBCC. Moreover, arterioles are preferable to venules whose flow is less stable and is more likely to be interrupted by rolling leukocytes. Arterioles are also preferable to smaller capillaries^16^, whose flow is even less robust (blood flow in small capillaries can even change direction), with the frequent presence of plasma gaps, which can lead to leukocyte detection artifacts.

To further improve the accuracy of image-based WBCCs, we average results from multiple vessels from the same participant (Fig. 2g). This approach significantly reduces the mean absolute percentage error (MAPE) from 45% to 27% when averaging measurements from 3-5 vessels, compared to using single-vessel data. This important finding suggests that averaging multiple vessels provides a more accurate assessment of systemic immune status, accounting for uneven leukocyte distribution in the microvasculature, which is determined by the branching order of individual vessels in the vascular tree^17^. In addition, factors like blood flow velocity and local hematocrit can orchestrate leukocyte margination^18,19^, which can impact their partitioning after bifurcation, and the overall distribution across the vasculature network.

The method described here represents an important advancement toward the noninvasive measurement of WBCC in humans, with the potential for substantial clinical impact, for example in monitoring critically ill patients in ICUs^20^, immunosuppressed cancer patients undergoing chemotherapy^21^, and preterm infants^2^, for whom the traditional blood draw approach poses high risks of secondary infections, anemia, and pain. The noninvasive aspect of mOBM also enables the continuous monitoring of the immune system, which is critical for timely clinical interventions in cases like sepsis, wherein every hour of delayed treatment reduces survival chances by 9%^22^. Another key application of mOBM is in preventive healthcare for the early detection and treatment of disease in the overwise considered healthy adult population. Indeed, WBC counts at the high end of the normal range have been linked to chronic systemic inflammation and to subclinical disease progression, especially among elderly and people with a higher risk of developing cardiovascular diseases, autoimmune diseases, cancer, type 2 diabetes, and depression among others^3^. Traditionally, monitoring such subclinical states would require repetitive blood draws throughout the day to capture fluctuations in leukocyte levels, a process that is problematic in routine healthcare due to the invasiveness, risk of infection, and potential for anemia, particularly in at-risk groups. Our method can bridge this gap and enable frequent WBCC measurements for the timely diagnosis and management of subclinical, developing diseases. Finally, the low cost and portability of the developed mOBM system make it particularly valuable in resource-poor settings (e.g., regions that lack infrastructure such as specialized equipment or advanced health care facilities.), allowing for simple noninvasive immune status assessment in the most difficult circumstances.

As a future development, we are working on developing a large FOV imaging system that will record multiple vessels in a single video acquisition. This will improve the accuracy of mOBM and decrease the imaging time of patients, which is currently between 30-50 minutes. Another key improvement involves the development of an automated machine-learning pipeline to enhance cell detection, accelerate image processing time, and classify different leukocyte subtypes (e.g., neutrophils vs. lymphocytes) based on cell size and granularity (Fig. 1d). Further miniaturization of mOBM will accelerate its integration into clinical practice and facilitate its use in neonates, ICU patients, and resource-poor settings.

## Materials and Methods

### Miniaturized oblique back-illumination microscope (mOBM)

The instrument illustrated in Figure 1a is the second generation of the recently outlined system (Bagramyan A and Lin CP., ‘‘*Miniaturized microscope for non-invasive imaging of leukocyte-endothelial interaction in human microcirculation’’*, Scientific Reports, 2023). The new design offers a more compact device, with a ≈35% reduction in length (from 18.7cm to 12.3cm), a ≈ 17% reduction in weight (from 0.157kg to 0.130kg), along with an expanded ≈320µmx260µm imaging field-of-view (FOV) essential for effective navigation and vessels selection within the lip’s microvascular network.

The optical layout of mOBM was designed in Zemax Optical Studio software (Supplementary Fig. 1a). To achieve a compact system, the number of components was reduced to four (Supplementary Fig. 1). The first component (1) is an aberration-corrected imaging GRIN lens assembly (Grintech, *GT-MO-080-032-ACR-VISNIR-08-20)* with a high numerical aperture (NA) of 0.75 (in water) that helps to maximize the optical resolution of mOBM. The second component (2) is a ≈¼ pitch relay GRIN lens (Edmund Optics, *#64-519*) that collimates the image rays before transmitting them to the third component (3), an achromat doublet lens (Edmund Optics, *#47-689*) with an effective focal length of 10 mm. This component (3) projects the magnified image onto the fourth component (4), a CMOS sensor (Basler, *daA1920-160um)* capable of fast image acquisition up to 1000 frames per second.

To generate oblique back-illumination, we use the epi-illumination configuration wherein the optical fiber (multimode, 1 mm core diameter) is positioned next to the imaging GRIN lens assembly (Supplementary Fig. 1c). The generated phase-gradient contrast (PGC) reveals fine morphological details (e.g., cell borders, intracellular granules) of blood cells (Fig. 1d). As imaging wavelength, we use 565 nm (Thorlabs, Solis-565D) that generates an absorption contrast between the red and white blood cells. The output power of the optical fiber was typically approximately 30mW, below the maximum permitted exposure limit of 34mW.

An important key element of mOBM is the imaging tip, designed with four channels: a central channel housing the miniaturized GRIN objective lens, an illumination channel holding the tip of the optical fiber, an irrigation channel to prevent the oral mucosa tissue from drying and to maintain immersion medium (water) in front of the GRIN lens, and a ring-shaped vacuum cavity to stabilize the imaging tissue. The instrument’s magnification was ≈ 13.45, yielding around 7.8 pixels/µm without binning and approximately 3.9 pixels/µm with 2×2 binning. The working distance (WD) of the imaging GRIN lens assembly (Supplementary Fig. 1a) was adjusted using motorized camera displacement along the optical axis. The mOBM’s mechanical housing, imaging tip, and oral mucosa apparatus (Fig. 1c) were designed in SolidWorks software (Supplementary Fig. 1b) and printed using a 3D laser printer (Formlabs 3B). All parts in contact with human tissue were printed from biocompatible materials (Formlabs, RS-F2-BMCL-01, RS-F2-BMBL-01).

### Oral Mucosa Apparatus for lip stabilization

To gently expose the lower lip’s microvasculature, we employed a universal oral mucosa apparatus developed in our laboratory (Bagramyan A and Lin CP., ‘‘*Miniaturized microscope for non-invasive imaging of leukocyte-endothelial interaction in human microcirculation’’*, Scientific Reports, 2023). The mechanical contours of the apparatus were tailored to conform to the lips’ morphology to minimize the high-pressure points that could potentially disrupt blood flow.

### Imaging of Healthy Participant

The imaging and data collection started after the participant (healthy volunteer) signed the IRB-approved consent form (MGH Protocol #:2021P003047). Initially, the participant was seated in front of the imaging system, and the chair and chin holder were adjusted accordingly. The lower lip was then gently unrolled using the oral mucosa apparatus, followed by an imaging session that typically lasted between 20 and 60 minutes. During imaging, the operator used a motorized XYZ actuator (mounted with mOBM) with micrometer resolution to navigate through the exposed lip tissue (Fig. 1b) for the identification and recording of the vessels of interest.

### Image acquisition parameters

Pylon camera software (Basler) was used for image acquisition. The acquisition frame rate was between 200-300 fps. Binning (2×2) was often used to increase the signal-noise ratio (SNR), reduce the exposure time (0.5 ms), and decrease the size of the recorded video files (.avi).

### Boltzmann Fit

The efficiency of WBC detection using mOBM is optimal when the cells are in focus and not masked by surrounding RBCs. In capillaries and small vessels, WBCs occupy the entire lumen, unobscured by RBCs (Fig. 1d, i-iv), and when in focus, nearly all WBCs within the sampled blood volume are detected. However, as the vessel diameter increases, the lumen cross-section can accommodate multiple cells, and WBCs may be outside the imaging plane or masked by RBCs that travel more superficially through the blood vessel (i.e. between the WBC and the microscope). In these situations, WBCs can be missed, leading to underestimation of WBCC (Supplementary Figure 2). The larger the diameter of the blood vessel, the higher the chance that WBCs will go undetected and the lower the WBCC measurement compared to the reference value (Supplementary Figure 2a).

To model the underestimation of WBCC detection with mOBM, a binary logistic regression model is represented by a Boltzmann function. This model is appropriate for our case, in which the binary cell detection (detected or not detected) is a function of blood vessel diameter. For small vessels, the Boltzmann function approaches 1 (Supplementary Figure 2a) due to efficient WBC detection. However, the fit curve gradually decreases as the instances of missed WBCCs increase in larger vessels. We use the fitted Boltzmann function to correct the corresponding raw WBCC measurements, accounting for leukocytes that were not detected within the sampled blood volume. Results are presented in Supplementary Figure 2.

### Measurements of vessel luminal diameter, blood flow velocity and WBC count

Automatic batch measurements of vessel luminal diameter, blood flow velocity and WBC count are made using custom software (*Supplementary Methods*). A region of interest (ROI) is defined with user assistance, and the software creates a coordinate system dictated by the shape of the ROI and allows time- and space-dependent measurements based on pixel intensity (*Supplementary Methods, Fig. 3*). To measure the vessel luminal diameter, a vessel lumen mask is drawn by locating pixels in the ROI with frequent and significant video-frame-to-video-frame pixel value changes. The vessel luminal diameter is the vessel-wall-to-vessel-wall distance of the mask (*Supplementary Methods, Fig. 3*). Blood flow velocity is measured using space-time diagrams, each constructed by rearranging pixels along a designated line that is parallel to the flow axis (*Supplementary Methods, Fig. 4*). RBC stripes in the diagrams are skeletonized into lines of weighted slope values, the statistics of which is reported as a function of time and space (*Supplementary Methods, Fig. 4*). WBC count is measured by counting the peaks in a background-corrected temporal trace of mean pixel values within a cell count window, the size and position of which are determined automatically by mapping the local contrast and vessel lumen diameters within the ROI. The threshold for peak finding is defined as 3.5x the scaled mean absolute deviation of the residual noise after background correction (*Supplementary Methods, Fig. 5-6*). WBCC is the WBC count divided by the blood flow volume, which is the multiple of blood flow velocity and vessel luminal area. The WBCC stability criterion is fulfilled by the ROI if the volume-binned cumulative WBCC values in the last ≥20nL of the ROI’s imaged flow volume falls within 5% of the final binned cumulative WBCC value (*Supplementary Methods, Fig. 7*).

## Data Availability

The data that support the findings of this study are available from the corresponding author upon reasonable request.

## Author contributions

A.B. conceived, designed, and built the system under the supervision of C.L.; A.B. performed the experiments. J.W. performed the data processing. A.B., J.W., K.M., C.A. and C.L. analyzed the data and wrote the manuscript.

## Competing interests

The authors declare no competing interests.

## Materials & Correspondence

Correspondence and requests for materials should be addressed to A.B.

## Acknowledgment

We would like to thank Dr. Jeffrey Gelfand for his generous help with human studies and Alice Chao for her contribution to manual cell counting. This work is supported in part by DOD grant FA9550-20-1-0063 and FA9550-23-1-0656, and by the Massachusetts General Hospital Executive Committee Fund for Medical Discovery and the SPIE Franz Hillenkamp Fellowship to A.B.

## Notes

### Competing Interest Statement

The authors have declared no competing interest.

### Funding Statement

This work is supported in part by DOD grants and by the Massachusetts General Hospital Executive Committee Fund for Medical Discovery and the SPIE Franz Hillenkamp Fellowship to A.B.

### Author Declarations

The experimental protocols related to human volunteers were performed in accordance with the guidelines and regulations of Massachusetts General Hospital. The study protocol (2021P003047) was approved by the Internal Review Board (IRB) of Massachusetts General Hospital. Informed consent was obtained from all subjects participating in our study.

